# Highly Sensitive Assays for detection of headache inducing neuropeptides, Pituitary adenylate cyclase-activating polypeptide (PACAP) and Calcitonin gene-related peptide (CGRP)

**DOI:** 10.1101/2025.03.04.25323177

**Authors:** Ourania Tzara, Josefine Nielsen Søderberg, Justyna Bahl, Emelie Andersson, Dina Silke Malling Damlund, Ib Vestergaard-Klewe, Mikkel Nors Harndahl, Allan Jensen, Ayodeji A. Asuni

## Abstract

**Background:** Migraine is one of the most disabling diseases that continues to pose a significant societal burden. Although there are now treatment options for people with migraine, it remains challenging to identify them as clinical features are diverse and complex, and there are no validated diagnostic or treatment prediction biomarkers. Identification is based on either diagnostic coding or the use of certain acute headache abortive treatments. However, socioeconomic disparities can contribute to under-diagnosis and under-treatment of migraine. Thus, efforts to find biomarkers to identify individuals with migraine and which variables could explain migraine-related chronification and disability are warranted. We aimed to investigate the levels of migraine inducing neuropeptides; calcitonin gene-related peptide (CGRP) and pituitary adenylate cyclase-activating polypeptide (PACAP) in peripheral blood samples as potential biomarkers of migraine.

**Methods:** We developed highly sensitive assays for CGRP and PACAP on the MSD S-PLEX assay platform and used them for bioanalysis of preclinical and clinical samples. Wildtype and neuropeptide challenged mice and rats were profiled using the developed assay. To follow-up, commercially obtained plasma samples from healthy controls and migraineurs were initially profiled. Subsequently, we profiled plasma samples from people with migraine (during and after a headache attack and healthy controls. Both MSD S-PLEX assays were transferred to Celerion where they were validated for analysis of clinical samples.

**Results:** Using the highly sensitive PACAP assay, we were able to reliably measure circulating levels of endogenous and administrated PACAP38in mouse and rat plasma. Additionally, using the highly sensitive CGRP assay, we were able to reliably measure circulating levels of endogenous and administrated CGRP in mouse and rat plasma. Furthermore, in the initial human samples, circulating CGRP and PACAP levels were not significantly different in healthy controls compared to people with migraine patients. However, ≥50% people with migraine showed increased circulating CGRP and PACAP levels during their attack period compared to post attack. Overall, people with migraine showed a 3 – 396% increase in one or both neuropeptides during their attack period compared to post attack. Circulating plasma CGRP and PACAP levels in healthy control subjects were consistent with previously measured levels.

**Conclusion:** Our highly sensitive PACAP and CGRP assays were successful in measuring circulating levels of endogenous PACAP38 and CGRP in mouse and rat plasma. Our highly sensitive PACAP and CGRP assays were qualified for measurement of human CGRP and PACAP in healthy control and migraine samples. Plasma CGRP and PACAP levels are elevated in migraineurs during an attack period, and the increased plasma neuropeptide levels during an attack may help the differentiation of migraineurs from non-Migraineurs, or amongst people with migraines to help identify the best treatment for each patient.

## INTRODUCTION

Migraine is a prevalent neurological disorder affecting nearly one billion individuals globally, and it is the leading cause of disability among all neurological conditions^1^. Many sufferers require preventive therapies to manage the frequency, severity, and duration of their migraine episodes^2^. Traditional preventive treatments, such as propranolol and topiramate are often abandoned due to insufficient efficacy or undesirable side effects. In recent years, scientific advancements and our understanding of migraine pathophysiology has introduced monoclonal antibodies and small molecule receptor antagonists that target calcitonin gene-related peptide (CGRP)^3^. Despite these innovations, a substantial number of people with migraine do not respond adequately to CGRP-targeted therapies, highlighting the urgent need for new treatments based on alternative mechanisms^4^.

Central to our understanding of migraine pathogenesis are the signaling molecules CGRP and pituitary adenylate cyclase-activating polypeptide (PACAP)^5^. CGRP, consisting of 37 amino acids, has two isoforms, αCGRP and βCGRP, with αCGRP predominantly expressed in somatosensory nerves and CNS neurons, while βCGRP is found mainly in enteric and motor neurons. In the trigeminal system, CGRP contributes to nitric oxide synthesis and sensitization of trigeminal nerves, promoting central sensitization. The role of CGRP in migraine is evidenced by elevated plasma levels during attacks and by the efficacy of anti-CGRP and anti-CGRP receptors targeting antibodies, as well as CGRP receptor antagonists^6–8^.

PACAP, a member of the vasoactive intestinal peptide (VIP)/glucagon/secretin family, shares significant sequence homology with VIP and acts via three G-Protein-coupled receptors: PAC1R, VIPR1, and VIPR2 ^9,10^. It is widely expressed in both central and peripheral nervous systems, including migraine-related structures like the trigeminal ganglia. PACAP can alter the phase of the circadian rhythm of neuronal activity during the daytime (resetting of the biological clock) ^11^. Specifically, PACAP was demonstrated in projections from the retina to the suprachiasmatic nucleus (SCN) and the intergeniculate leaflet (IGL), two central sites that regulate the circadian system, and in the SCN^12,13^. PACAP was found to reset the phase of the biological clock through a PACAP-R1 receptor via a cAMP signaling pathway ^11^.

Experimental studies have shown that intravenous infusion of PACAP induces migraine-like attacks in individuals with a history of migraine^14^. The plasma levels of PACAP-38 rise during migraine attacks ^8,15,16^, suggesting an association between the plasma PACAP-38 levels and migraine episodes. This also suggests that blocking PACAP signaling might constitute a promising drug target for migraine prevention. This led to the development of Lu AG09222, a humanized mAb that potently and selectively binds and neutralizes PACAP, resulting in inhibition of PACAP signaling. Animal experiments have demonstrated that Lu AG09222 prevents neurogenic inflammation and vasodilation, which are markers of migraine in rodent models^17^, and a phase 1 trial showed it could inhibit PACAP-induced dilation of cranial arteries in humans^18^. No currently approved preventive treatment for migraines fully prevents the onset of attacks. Many patients experience inadequate improvement or face intolerable side effects, which significantly affect their productivity and quality of life ^19,20^.

Given the significant roles of CGRP and PACAP in migraine pathophysiology^21^, accurately measuring their levels in biological samples may be crucial for understanding their roles and continued development of related targeted therapies^22–24^. This manuscript focuses on the development and validation of highly sensitive and specific ligand binding immunoassays on the Mesoscale Discovery Platform (MSD) for CGRP and PACAP, enabling precise quantification in plasma and tissue extracts to support further research and therapeutic advances.

## MATERIALS AND METHODS

### Animal

#### Mice

Mice used in this study were purchased from Taconic (Denmark). Briefly, wildtype C57BL/6J mice were used. Mice were housed in isolated ventilated cages under 12:12 light/dark cycles with food (Altromin 1324; Altromin Spezialfutter, Lage, Germany) and water *ad libitum*. Room humidity and temperature were 55%±5% and 21±2°C, respectively. Mice were brought to the euthanization room immediately before euthanization to avoid acute stress induced by termination. Mice were euthanized by awake decapitation and blood was collected in 0.5 ml Microvette® EDTA K3E tubes (Sarstedt, Germany), centrifuged at 3600g for 15 min, 4°C to collect plasma. All tissue samples were snap frozen on dry ice and stored at −80°C until use.

#### PACAP-challenged mice

10 mice (n= 5 each) were dosed with vehicle or PACAP38 (0.6 mg/kg). Mice were brought to the euthanization room immediately before euthanization to avoid acute stress induced by termination. Mice were euthanized by awake decapitation and blood was collected into cooled K3EDTA-coated tubes, centrifuged for 10 min @ 3000 g, 4. Recovered plasma was aliquoted into micronic tubes for neuropeptide analysis. 100 ul in micronic tube plus a DPP-IV inhibitor (2 ul) and the remaining was stored in micronic tube minus the inhibitor until use.

#### Rats

Sprague-Dawley rats used in this study were purchased from Charles River (Germany). Briefly, wildtype SD rats were used. Rats were housed in isolated ventilated cages under 12:12 light/dark cycles with food (Altromin 1324; Altromin Spezialfutter, Lage, Germany) and water ad libitum. Room humidity and temperature were 55%±5% and 21±2°C, respectively.

#### PACAP-challenged rats

10 rats (n= 5 each) were dosed with vehicle or PACAP38 (0.6 mg/kg). Rats were brought to the euthanization room immediately before euthanization to avoid acute stress induced by termination. Rats were euthanized by awake decapitation and blood was collected into cooled K3EDTA-coated tubes, centrifuged for 10 min @ 3000 g, 4. Recovered plasma was aliquoted into micronic tubes for neuropeptide analysis. 150 ul in micronic tube plus a DPP-IV inhibitor (3 ul) and the remaining was stored in micronic tube minus the inhibitor until use.

All animal experiments were in accordance with the European Communities Council Directive no. 86/609, the directives of the Danish National Committee on Animal Research Ethics, and Danish legislation on experimental animals (license no. 2014-15-0201-00339 C01 and C05).

### Human Plasma Sample Preparation

Individual control and disease-state human plasma were acquired from Cureline (USA). Cureline coordinated the prospective sample collection under institutional review board (IRB) ethical approval and with informed donor consent (IRB protocol CU-606-4797). The samples include: 1) Patient screening and selection according to the specified criteria. 2) Specimen collection and preparation according to the protocol. The samples were deidentified (Pseudonymized) under Cureline SOP CTBO.003 titled: Biospecimen deidentification, labeling and tracking, issued June 22, 2010, revised March 22, 2012.

#### Plasma (1-2 ml), frozen

Whole blood is collected in K2EDTA tubes (BD, lavender top, catalog# 366643) and processed as the manufacturer directs within 2 (two) hours after collection. Aliquots (1 ml) were quickly frozen, stored at −80 °C for subsequent downstream bioanalysis.

#### P800 tubes plasma

Whole blood is collected in BD™ P800 Blood Collection System https://www.bdbiosciences.com/us/applications/blood-collection/cell-biomarker-preservation/bdtrade-p800-blood-collection-system/p/366420 or 366421) and processed as follows. BD™ P800 is an evacuated blood collection tube containing a proprietary cocktail of protease, esterase, and DPP-IV inhibitors which provides immediate protection of bioactive peptides from degradation in plasma. Blood was drawn into a vacutainer tube(s) containing K2EDTA. The vacutainer tubes were inverted carefully 10 times to mix blood and anticoagulant and store at room temperature until centrifugation. The samples were centrifuged immediately in a swing type centrifuge, for 10 minutes at 1000-2000 RCF at room temperature. 3 layers were formed, and supernatant (plasma) was collected, aliquoted and stored at −80 °C for subsequent downstream bioanalysis.

### 3 sets of human samples were collected for the initial feasibility studies

Set 1. Number of cases: N = 12 Migraine (Baseline (No headache attack)) and 12 age-matched Normal controls (age-matched +/- 5 yrs.). Both, 1 ml K2EDTA plasma and 1 ml P800 plasma were collected.

Set 2. Number of cases: N = 24 healthy donors (age-matched +/- 5 yrs.). Both, 1 ml K2EDTA plasma and 1 ml P800 plasma were collected.

Set 3: Number of cases: N = 12 Migraine (Baseline (No headache attack) T1, Headache attack T2) and 12 age-matched Normal controls age-matched +/- 5 yrs.). Female to Male ratio −13/7 Collection time points: T1-During Migraine or stress headaches attack. T2-Before or after the Migraine or stress headache attack has subsided.

### Statistical Analysis

All statistical analyses were carried out using GraphPad Prism 10.0 (GraphPad Software Inc., La Jolla, CA, USA). Data analysis was carried out using Excel 2016 (Microsoft Corporation, Redmond, WA, USA).

## Neuropeptide Assay Development

### Reagents and Buffers for PACAP assay

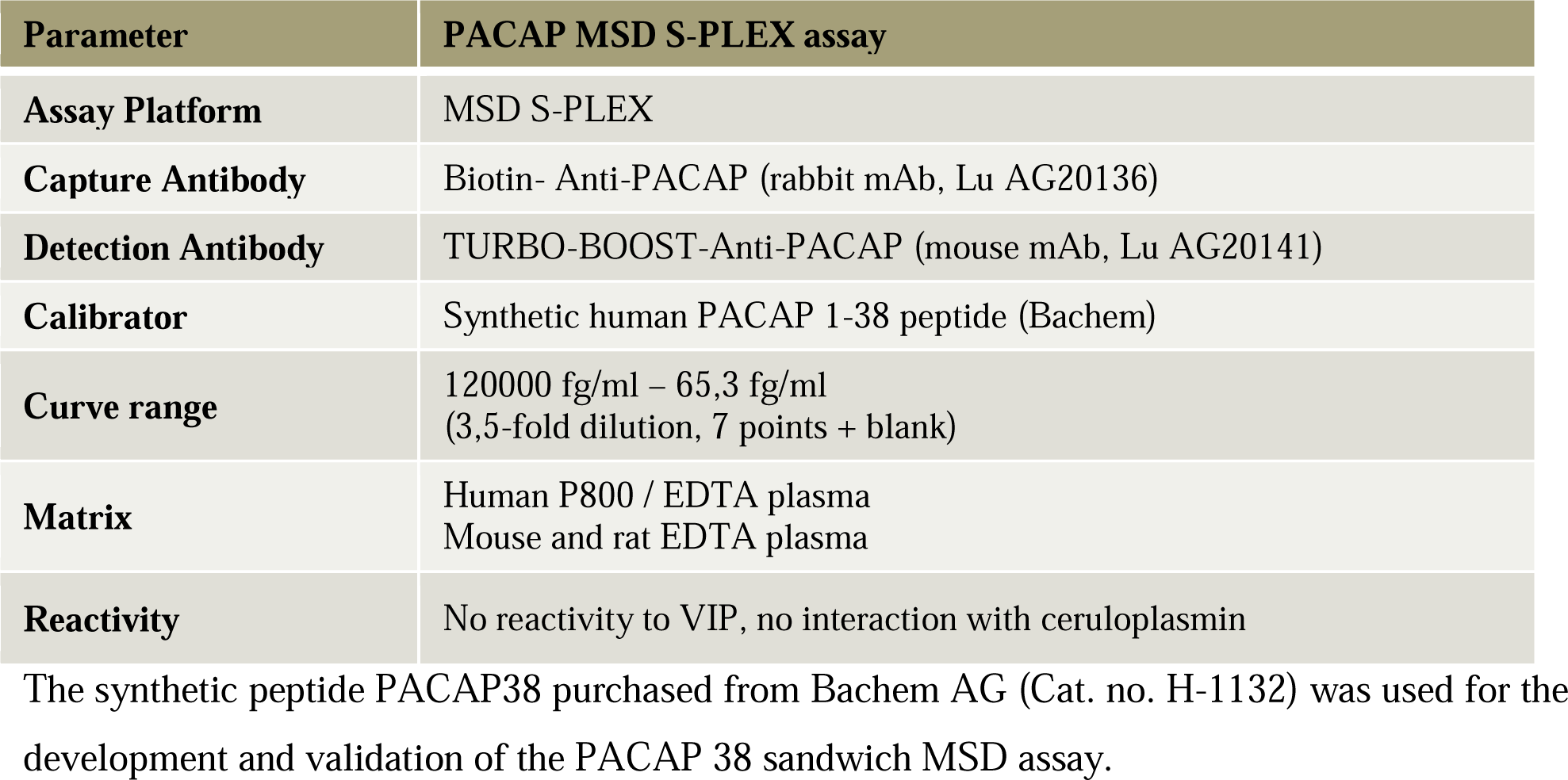

| Parameter | PACAP MSD S-PLEX assay |
| --- | --- |
| Assay Platform | MSD S-PLEX |
| Capture Antibody | Biotin- Anti-PACAP (rabbit mAb, Lu AG20136) |
| Detection Antibody | TURBO-BOOST-Anti-PACAP (mouse mAb, Lu AG20141) |
| Calibrator | Synthetic human PACAP 1-38 peptide (Bachem) |
| Curve range | 120000 fg/ml – 65,3 fg/ml<br>(3,5-fold dilution, 7 points + blank) |
| Matrix | Human P800 / EDTA plasma<br>Mouse and rat EDTA plasma |
| Reactivity | No reactivity to VIP, no interaction with ceruloplasmin |

The synthetic peptide PACAP38 purchased from Bachem AG (Cat. no. H-1132) was used for the development and validation of the PACAP 38 sandwich MSD assay.

### Antibodies for the PACAP assay

The PACAP assay is based on two proprietary antibodies. The biotinylated monoclonal anti-PACAP antibody (Biotin-Anti-PACAP (rabbit mAb, Lu AG20136)), which recognizes both PACAP27 and PACAP38, was used as a capture antibody. Another anti-PACAP antibody (TURBO-BOOST-Anti-PACAP (mouse mAb, Lu AG20141) that also binds PACAP, was used as a detection antibody.

### Principle of the PACAP assay

The MSD 96-well S-PLEX Human/Mouse/Rat PACAP assay is a sandwich immunoassay. A biotinylated capture antibody is coupled to streptavidin on the spot of the S-PLEX plate. Analytes in the sample bind to the capture antibody, and detection antibody conjugated with the TURBO-BOOST label binds to the analyte to complete the sandwich immunoassay (Figure 1). Once the sandwich immunoassay is complete, the TURBO-BOOST labeled detection antibody is enhanced by the addition of a TURBO-TAG label. Finally, the user adds MSD read buffer that creates the chemical environment for electrochemiluminescence and loads the plate onto an MSD instrument where a voltage applied to the plate electrodes causes the captured labels to emit light. The instrument measures the intensity of emitted light, which is proportional to the amount of PACAP present in the sample and provides a quantitative measure of the analyte. The concentrations of PACAP in samples were interpolated against a standard curve made up of reference standard using a 4 PL fit (MSD workbench). The assay was used in-house for routine bioanalysis of preclinical samples, and subsequently validated for clinical use at Celerion.

**Figure 1.**
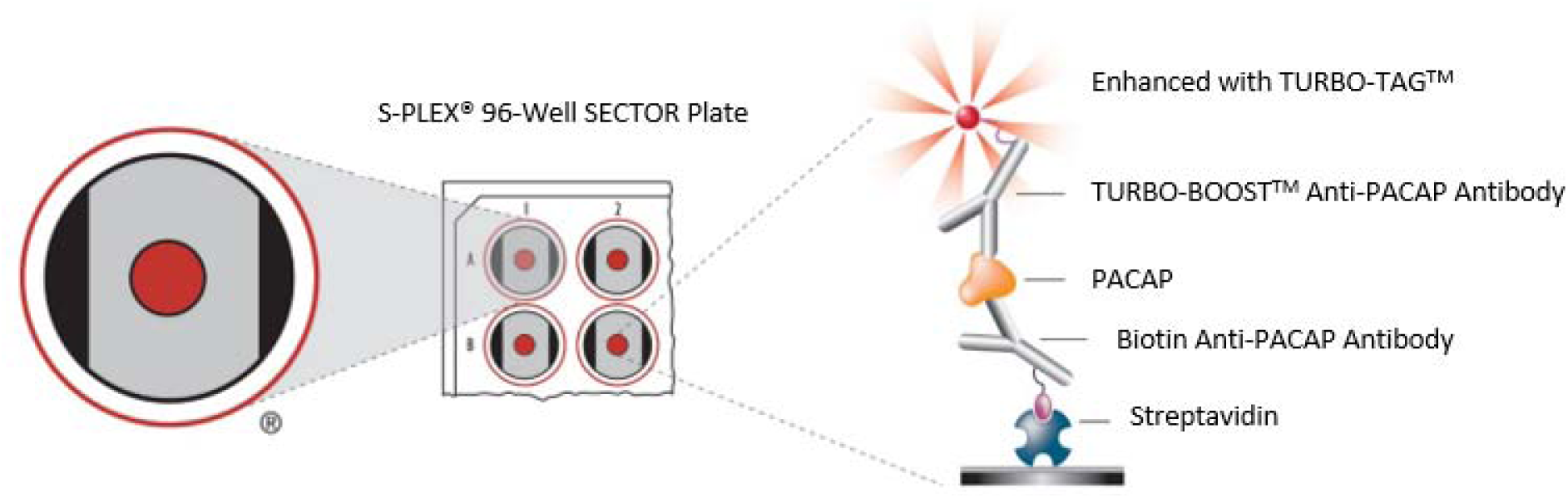
Principle of MSD 96-well S-PLEX Human/Mouse/Rat PACAP assay.

### Reagents and Buffers for CGRP assay

| Parameter | CGRP MSD S-PLEX assay |
| --- | --- |
| Assay Platform | MSD S-PLEX |
| Capture Antibody | Biotin-Anti-CGRP (Lu AG14079) |
| Detection Antibody | TURBO-BOOST-Anti-CGRP (Bioporto Diagnostics, ABS-026-05) |
| Calibrator | Human: Synthetic human aCGRP peptide (Bachem)<br>Mouse/rat: Synthetic mouse/rat aCGRP peptide (Bachem) |
| Curve range | Human:100000 fg/ml – 24,4 fg/ml (4-fold dilution, 7 points + blank)<br>Mouse/rat: 22000 fg/ml – 30 fg/ml (3-fold dilution, 7 points + blank) |
| Matrix | Human P800 / EDTA plasma<br>Mouse, rat EDTA plasma |
| Reactivity | No reactivity to PACAP |

### Antibodies for the CGRP assay

The CGRP assay is based on two proprietary antibodies. The biotinylated monoclonal anti-CGRP antibody (Biotin-Anti-CGRP (Lu AG14079)), which recognizes CGRP, was used as a capture antibody. Another anti-CGRP antibody (TURBO-BOOST-Anti-CGRP (mouse mAb, Bioporto Diagnostics, ABS-026-05) that also binds CGRP, was used as a detection antibody.

### Principle of the CGRP assay

The MSD 96-well S-PLEX Human and Mouse/Rat CGRP assays are sandwich immunoassays. A biotinylated capture antibody is coupled to streptavidin on the spot of the S-PLEX plate. Analytes in the sample bind to the capture antibody, and detection antibody conjugated with the TURBO-BOOST label binds to the analyte to complete the sandwich immunoassay (see Figure 2). Once the sandwich immunoassay is complete, the TURBO-BOOST labeled detection antibody is enhanced by the addition of a TURBO-TAG label. Finally, the user adds MSD read buffer that creates the chemical environment for electrochemiluminescence and loads the plate onto an MSD instrument where a voltage applied to the plate electrodes causes the captured labels to emit light. The instrument measures the intensity of emitted light, which is proportional to the amount of CGRP present in the sample and provides a quantitative measure of the analyte. The concentrations of CGRP in samples were interpolated against a standard curve made up of reference standard using a 4 PL fit (MSD workbench).

**Figure 2.**
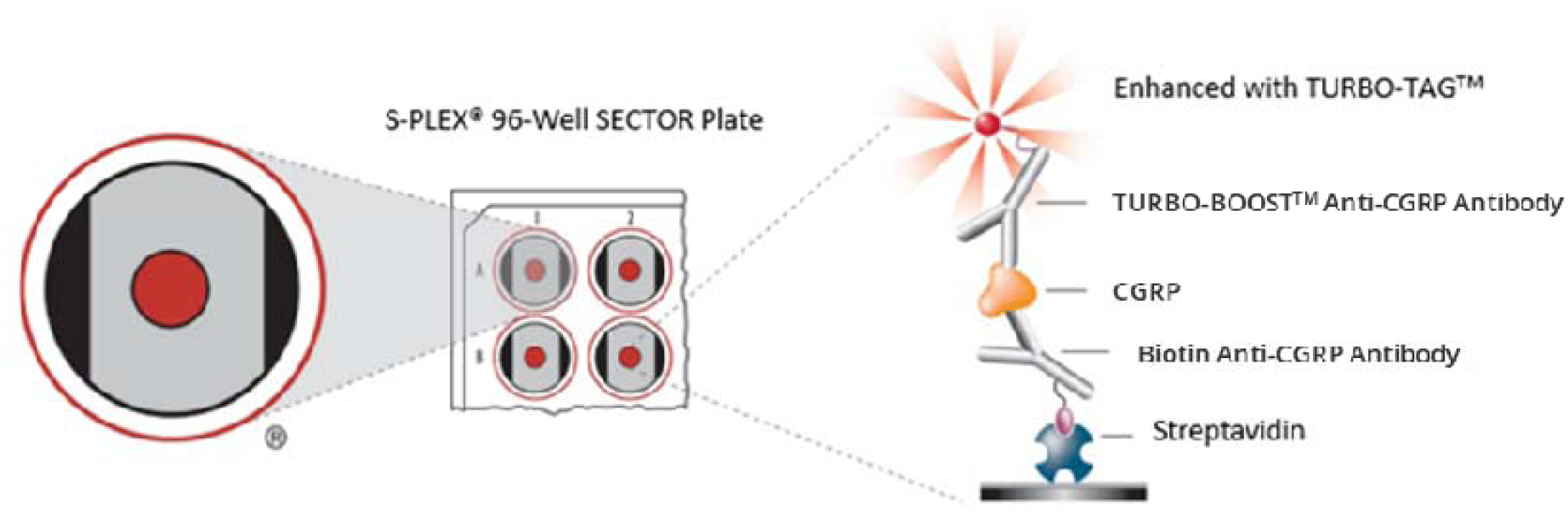
Principle of MSD 96-well S-PLEX Human/Mouse/Rat CGRP assay.

The assay was used in-house for routine bioanalysis of preclinical samples and subsequently validated for clinical use at Celerion.

Human S-PLEX PACAP assay method validation.

**Human S-PLEX PACAP assay method validation**
| Method Validation Summary |  |  |  |
| --- | --- | --- | --- |
| Analyte | PACAP |  |  |
| Matrix | Human EDTA plasma |  |  |
| Method SOP | SM3-518 |  |  |
| Assay method | ECLIA -S-PLEX |  |  |
| Assay volume | 120 µL |  |  |
| Minimum required dilution MRD | 2 |  |  |
| Analytical range | 50-80 000 fg/mL |  |  |
| Regression model | 4-PL, weighting 1/y<sup>2</sup> |  |  |
| Precision and accuracy / sensitivity | Precision<br>(%CV) | Bias<br>(%) | Total Error<br>(%) |
| Between-run: |  |  |  |
| LLOQ-QC (50.0 fg/mL) | 15.9 | -15.6 | 31.5 |
| LQC (150 fg/mL) | 22.0 | 0.0 | 22.0 |
| MQC (2'000 fg/mL) | 7.6 | 14.5 | 22.1 |
| HQC (60'000 fg/mL) | 5.6 | 10.3 | 16.0 |
| ULQC (80'000 fg/mL) | 10.9 | -5.1 | 16.1 |
| Selectivity (healthy population) | All 10 individual matrix lots that were investigated were within acceptance. |  |  |
| Long-term (frozen) stability in plasma | Long term stability assessment ongoing. |  |  |
| Freeze-thaw and short-term (bench-top) stability in plasma | Demonstrated for 3 and 6 freeze/thaw cycles at -80 °C to room temperature in EDTA P800 polypropylene tubes with a longest single thaw period of 3 hours and a cumulative thaw period of 10 hours (in 3 freeze/thaw cycles) and of 20 hours (in 6 freeze/thaw cycles).<br>Short term stability demonstrated for 69 days. |  |  |
| Stability for solutions | Demonstrated in polypropylene tubes for:<br>WS1: 87 days at -80 °C.<br>STD 3 and STD 9: 80 days at -80 °C.<br>SL needs to be used fresh. |  |  |
| Stability during sample collection and handling | Demonstrated in EDTA tubes.<br>Demonstrated in P800 tubes. |  |  |

#### Test items

The test item is identical to the reference item.

*Reference item*

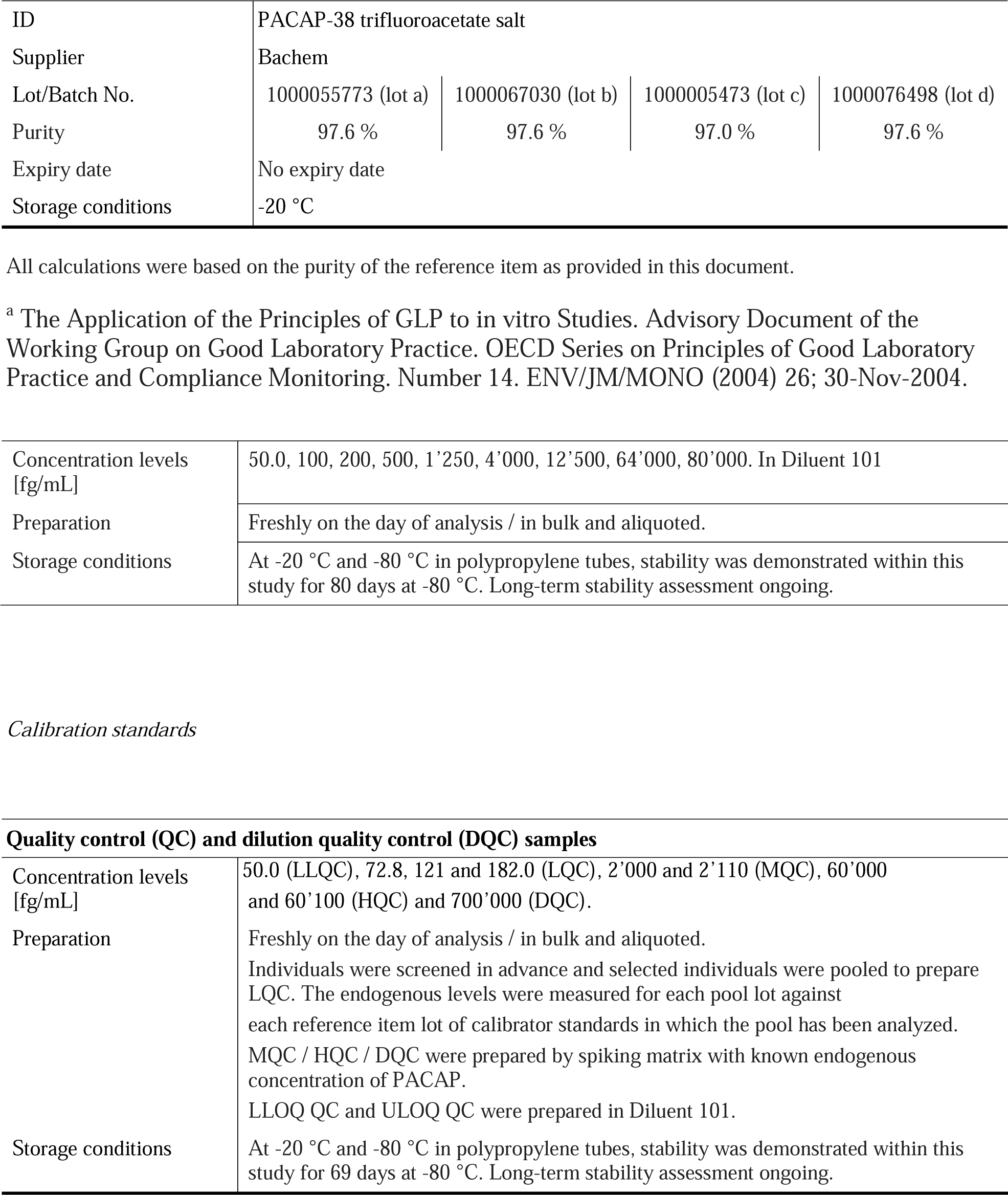

|  |  |  |  |  |
| --- | --- | --- | --- | --- |
| ID | PACAP-38 trifluoroacetate salt |  |  |  |
| Supplier | Bachem |  |  |  |
| Lot/Batch No. | 1000055773 (lot a) | 1000067030 (lot b) | 1000005473 (lot c) | 1000076498 (lot d) |
| Purity | 97.6 % | 97.6 % | 97.0 % | 97.6 % |
| Expiry date | No expiry date |  |  |  |
| Storage conditions | -20 °C |  |  |  |

|  |  |
| --- | --- |
| Concentration levels [fg/mL] | 50.0, 100, 200, 500, 1'250, 4'000, 12'500, 64'000, 80'000. In Diluent 101 |
| Preparation | Freshly on the day of analysis / in bulk and aliquoted. |
| Storage conditions | At -20 °C and -80 °C in polypropylene tubes, stability was demonstrated within this study for 80 days at -80 °C. Long-term stability assessment ongoing. |

**Quality control (QC) and dilution quality control (DQC) samples**
|  |  |
| --- | --- |
| Concentration levels [fg/mL] | 50.0 (LLQC), 72.8, 121 and 182.0 (LQC), 2'000 and 2'110 (MQC), 60'000 and 60'100 (HQC) and 700'000 (DQC). |
| Preparation | Freshly on the day of analysis / in bulk and aliquoted.<br>Individuals were screened in advance and selected individuals were pooled to prepare LQC. The endogenous levels were measured for each pool lot against each reference item lot of calibrator standards in which the pool has been analyzed. MQC / HQC / DQC were prepared by spiking matrix with known endogenous concentration of PACAP.<br>LLOQ QC and ULOQ QC were prepared in Diluent 101. |
| Storage conditions | At -20 °C and -80 °C in polypropylene tubes, stability was demonstrated within this study for 69 days at -80 °C. Long-term stability assessment ongoing. |

#### Analytical run acceptance criteria

An analytical run was acceptable if the following criteria were met: at least 75% of non-anchor point calibration standards (and at least six) meet acceptance criteria: CV of replicate responses ≤ 30.0% (≤ 30.0% at the LLOQ and ULOQ) and mean back-calculated concentration within ± 30.0% (± 30.0% at the LLOQ and ULOQ) of their nominal concentration, at least two-thirds of the QC samples and at least 50% at each concentration level meet acceptance criteria: CV of replicate calculated concentrations ≤ 30.0% and mean back-calculated concentration within ± 30.0% of their nominal concentration. However, the dedicated accuracy and precision (A&P) runs have no QC acceptance criteria and the blank response is below the LLOQ response. QC and/or validation samples out of acceptance are flagged and their values documented in the method validation report accordingly. QC and/or validation samples failing to meet criteria for analytical reasons were excluded from evaluation and can be repeated. However, QC samples used to evaluate A&P are only excluded if an assignable technical reason is documented. The QC data and statistics for non-A&P runs are reported independently from the QC samples used to evaluate the precision and accuracy of the method.

#### Accuracy and precision (within-run and between-run)

The accuracy and precision of the method were determined in eight independent analytical runs performed on three different days. Each of the runs included six replicate QC samples at five concentration levels (LLOQ-QC, LQC, MQC, HQC, ULOQ-QC). For four of the runs, formerly prepared and stored calibration standards and QC samples were used. For two of the runs, calibration standards were freshly prepared. One of the runs contained a total number of samples that reflects the maximum size of an anticipated sample analysis run (full 96-well plate). All A&P runs met acceptance criteria for all runs.

#### Selectivity (healthy population)

Selectivity was determined by analysing 10 healthy individual human plasma lots as blank sample (un-spiked) and spiked with 1’000 fg/mL. Each individual lot was analyzed in 1 replicate. All lots met acceptance criteria.

### Stabilities

#### Freeze-thaw and short-term (bench-top) stability in matrix

Freeze-thaw and short-term stability human plasma (EDTA P800) was evaluated using six replicates, from at least 3 aliquots of LQC (endogenous concentration) and spiked samples at two concentration levels (HQC and DQC) in polypropylene tubes exposed to the following conditions:

- Thawing and storage at ambient temperature for a longest thaw period of 3.9 hours and a cumulative thaw period of 20 hours (6 freeze/thaw cycles) and for a longest thaw period of 3.9 hours and a cumulative thaw period of 10 hours (3 freeze/thaw cycles).
- Number of freeze-thaw cycles: 3 and 6.

Stability samples were analyzed against freshly prepared calibration standards. The initial freezing period was a minimum of 24 hours, and for each subsequent freeze/thaw cycle the stability samples were stored frozen for a minimum of 12 hours followed by a thawing under appropriate conditions for typically 3 hours.

#### Long-term (frozen) stability in matrix

Long-term stability in human plasma (EDTA P800) was evaluated using six replicates, from at least 3 aliquots of LQC (endogenous concentration) and spiked samples at HQC level. Stability samples stored at −80 °C for 69 days in polypropylene tubes were analyzed against freshly prepared calibration standards. Stability was demonstrated.

Further long-term stability assessment will be assessed in study CA34093-02.

#### Stability for solutions

Stability of the stock solution (1’000 μg/mL in Millipore water), working solution (10’000 ng/mL in Diluent 100), STD 3 (100 fg/mL), and STD 9 (64’000 fg/mL) were evaluated using six replicates, from at least 3 aliquots, in polypropylene tubes. Stability samples stored at −80 °C for 38 and 80 days (STD 3 and STD 9), 46, 53 and 87 days (WS 1) and for 26, 28 and 34 days (SL) were analyzed against freshly prepared calibration standards. The stored replicates were compared with the replicates from a freshly prepared working solution. Stability was demonstrated for STD3 and STD 9 for 80 days, and for WS 1 for 87 days.

#### Stability during sample collection and handling (Plasma)

Short-term stability of the analyte during sample collection and handling in human whole blood sample was evaluated blank and at MQC level (2’000 fg/mL). The samples were incubated in regular K_2_EDTA tubes and P800 tubes at room temperature for 0 (control samples at T0h) and 2 hours (stability samples at T2h) to mimic the anticipated collection conditions. At each time point, blood was processed and was divided into appropriate aliquots for analysis. Whole blood stability was evaluated by processing six replicates of blank and MQC level samples from each time point. No significant interference was observed in the whole blood. Stability was demonstrated in polypropylene tubes.

### CGRP assay method validation

| **Method Validation Summary** |  |  |  |
| --- | --- | --- | --- |
| Analyte | CGRP (Calcitonin Gene Related Peptide) |  |  |
| Matrix | Human plasma (EDTA) with/without P800 |  |  |
| Method SOP | SM3-523 |  |  |
| Assay method | ECLIA S-PLEX |  |  |
| Assay volume | 25 µL |  |  |
| Minimum required dilution MRD | 8 |  |  |
| Analytical range | 200 - 80'000 fg/mL (0.200 pg/mL to 80.0 pg/mL) |  |  |
| Regression model | 4-PL Marquardt, weighting 1/y<sup>2</sup> |  |  |
| Precision and accuracy | Precision<br>(%CV) | Bias<br>(%) | Total Error<br>(%) |
| Between-run: |  |  |  |
| LLOQ-QC (0.200 pg/mL) | **37.5** | -17.8 | **55.3** |
| LQC (0.267 pg/mL) | 16.3 | 4.9 | 21.1 |
| MQC (1.23 pg/mL) | 19.3 | 0.0 | 19.3 |
| HQC (56.0 pg/mL) | 16.6 | 12.0 | 28.6 |
| ULOQ-QC (80.0 pg/mL) | 8.0 | 1.0 | 9.0 |
| Selectivity (healthy population) and matrix effect | 10 out of 10 K<sub>2</sub> EDTA P800 individual matrix lots that were investigated were within acceptance.<br>1/1 hemolyzed and 1/1 lipemic K<sub>2</sub> EDTA P800 sample met acceptance criteria |  |  |
| Freeze-thaw and short-term (bench-top) stability of quality control samples | LQC - K<sub>2</sub> EDTA P800 at -20°C: 6 freeze/thaw cycles with a single longest thaw time of 4.5 hours at room temperature, a total thaw time of 20.4 hours and a single longest freezing time of 7 days.<br>LQC - K<sub>2</sub> EDTA at -80°C: 3 freeze/thaw cycles with a single longest thaw time of 4.5 hours at room temperature, a total thaw time of 11.7 hours and a single longest freezing time of 22 days.<br>HQC - Pierce Protein-Free T20 (TBS) Blocking Buffer at -80°C: 6 freeze/thaw cycles with a single longest thaw time of 5.1 hours at room temperature, a total thaw time of 22.1 hours and a single longest freezing time of 7 days. |  |  |
| Stability for solutions | At 1.00 mg/mL (SL) in DMSO, 10.0 µg/mL (WS1) and 0.500 pg/mL (STD 7) in Pierce Protein Free T20 (TBS) Blocking Buffer:<br>SL: 28 days at -80°C.<br>WS1: 2 freeze/thaw cycles with a single longest thaw time of 5.4 hours at room temperature, a total thaw time of 8.5 hours and a single longest freezing time of 27 days.<br>STD 7: 1 freeze/thaw cycles with a thaw time of 4.6 hours at room temperature and a freezing time of 50 days. |  |  |
| Stability during sample collection and handling | No significant interference was observed for samples prepared with K<sub>2</sub> EDTA or K<sub>2</sub> EDTA P800 in whole blood. |  |  |

#### Acceptance criteria for blank matrix

Prior to method validation, blank matrices were screened to determine the basal level of CGRP.

#### Analytical run acceptance criteria

An analytical run was acceptable if the following criteria were met:

- at least 75% of non-anchor point calibration standards (and at least six) meet acceptance criteria: CV of replicate responses ≤ 30.0% (≤ 30.0% at the LLOQ and ULOQ) and mean back-calculated concentration within ± 30.0% (± 30.0% at the LLOQ and ULOQ) of their nominal concentration,
- at least two-thirds of the QC samples and at least 50% at each concentration level meet acceptance criteria: CV of replicate calculated concentrations ≤ 30.0% and mean back-calculated concentration within ± 30.0% of their nominal concentration. However, the dedicated A&P runs have no QC acceptance criteria.
- the blank response is below the LLOQ response.

QC and/or validation samples out of acceptance are flagged and their values documented in the method validation report accordingly. QC and/or validation samples failing to meet criteria for analytical reasons were excluded from evaluation and can be repeated. However, QC samples used to evaluate A&P are only excluded if an assignable technical reason is documented. The QC data and statistics for non-A&P runs are reported independently from the QC samples used to evaluate the precision and accuracy of the method.

#### Accuracy and precision (within-run and between-run)

The accuracy and precision of the method at LQC and MQC (in K_2_ EDTA matrices), HQC and ULOQ-QC levels were determined in 6 independent analytical runs performed on four different days. Each of the runs included six replicate QC samples at the four concentration levels. For three runs, formerly prepared and stored calibration standards and QC samples were used. For the other 4 runs, calibration standards and QC samples were freshly prepared. Run 25 contained a total number of samples that reflects the maximum size of an anticipated sample analysis run (full 96-well plate). The accuracy and precision of the method at LLOQ-QC level (0.200 pg/mL) were determined in 10 independent analytical runs performed on six different days and each of the runs included six replicates. For four runs, formerly prepared and stored calibration standards and QC samples were used. In seven runs, calibration standards and QC samples were freshly prepared.

The acceptance criteria for accuracy and precision (within-run and between-run) are:

- Accuracy: the mean concentration should be within ± 30. % of the nominal values for each QC level.
- Precision: the CV should not exceed 30.0 % for each QC level (± 30.0% for LLOQ-QC and ULOQ-QC).
- Total error: The total error for each QC is ≤ 30.0 % (≤ 50.0 % for LLOQ QC and ULOQ-QC).

In total the within-run and between-run accuracy and precision were determined over 6 runs for LQC, MQC, HQC and ULOQ-QC, and over 10 runs for LLOQ-QC and met acceptance criteria for inter-run accuracy and precision all levels except for LLOQ-QC

#### Selectivity (healthy population)

Selectivity was determined by analysing 15 K_2_ EDTA and 10 K_2_ EDTA P800 healthy individual human plasma lots, and one haemolyzed and one lipemic matrix lot in each set-up, as blank sample (un-spiked) and spiked with 0.200 pg/mL of reference item. Each individual lot was analyzed in 1 replicate.

The acceptance criteria for selectivity are:

- The measured concentration should be within ± 30.0% of the nominal values for each level when compared to the blank concentration.
- At least 80 % of the tested matrices must meet the acceptance criteria.

For samples prepared in K_2_ EDTA P800 matrix, 10 out of 10 healthy individual matrix lots were within acceptance criteria, also the haemolyzed and lipemic samples met criteria.

### Stabilities

#### Freeze-thaw and short-term (bench-top) stability of quality control samples

Freeze-thaw and short-term stability of human plasma was evaluated using six replicates, from at least 3 aliquots spiked at HQC prepared with Pierce Protein-Free T20 (TBS) Blocking Buffer and LQC prepared with K_2_ EDTA or with K_2_ EDTA P800 matrices in polypropylene tubes.

The acceptance criteria for freeze-thaw and short-term (bench-top) stability are:

- Accuracy: The mean concentration of the stability samples should be within ± 30.0% of the nominal value.
- Precision: The CV of the stability samples should not exceed 30.0%.
- A minimum of 2/3 of the replicates must have results that can be evaluated.

Stability was demonstrated for the following conditions:

For HQC samples

- Thawing and storage at room temperature for a longest thaw period of 5.1 hours and a cumulative thaw period of 22.1 hours,
- Freezing at −80°C for a longest freezing time of 7 days.
- Number of freeze-thaw cycles: 6.

For LQC - K_2_ EDTA samples:

- Thawing and storage at room temperature for the longest thaw period of 4.5 hours and a cumulative thaw period of 11.7 hours,
- Freezing at −80°C for a longest freezing time of 22 days (based on the last basal level assessment run).
- Number of freeze-thaw cycles: 3.

For LQC - K_2_ EDTA P800 samples:

- Thawing and storage at room temperature for a longest thaw period of 4.5 hours and a cumulative thaw period of 20.4 hours,
- Freezing at −80°C for a longest freezing time of 7 days (based on the last basal level assessment run),
- Number of freeze-thaw cycles: 6.

Stability samples were analysed against freshly prepared calibration standards.

#### Long-term (frozen) stability in matrix

Long-term stability of CGRP is ongoing.

#### Stability for solutions

Stability of the stock solution SL (1.00 mg/mL) in DMSO and working solution WS1 (10.0 µg/mL), STD 8 (0.200 pg/mL) and STD 7 (0.500 pg/mL) in Pierce Protein Free T20 (TBS) Blocking Buffer were evaluated using six replicates, from at least 3 aliquots, in polypropylene tubes.

The acceptance criteria for stability for solutions are:

- Accuracy: The mean concentration of the stability samples should be within ± 30.0% of the nominal value.
- Precision: The CV of the stability samples should not exceed 30.0%.
- A minimum of 2/3 of the replicates must have results that can be evaluated.

Stability was demonstrated for the following conditions:

For SL samples:

- Freezing at −80°C for 28 days.
- Number of freeze-thaw cycles: 1.

For WS1 samples:

- Thawing and storage at room temperature for a longest thaw period of 5.4 hours and a cumulative thaw period of 8.5 hours,
- Freezing at −80°C for a longest freezing time of 27 days.
- Number of freeze-thaw cycles: 2.

For STD 7 samples: Thawing and storage at room temperature for a longest thaw period of 4.6 hours,

- Freezing at −80°C for a longest freezing time of 50 days,
- Number of freeze-thaw cycles: 1.

Stability samples were analysed against freshly prepared calibration standards.

#### Stability during sample collection and handling (Plasma)

Short-term stability of the analyte during sample collection and handling in fresh human whole blood was evaluated blank and at MQC level prepared with K_2_ EDTA (0.884 and 1.11 pg/mL) or with K_2_ EDTA P800 (1.12 pg/mL). The samples were incubated in polypropylene tubes at room temperature for 0 (control samples at T0h) and 2 hours (stability samples at T2h) to mimic the anticipated collection conditions. At each time point, blood was processed and was divided into appropriate aliquots for analysis. Whole blood stability was evaluated by processing six replicates of blank and MQC level samples from each time point.

The acceptance criteria for stability during sample collection and handling are:

- Accuracy (% of T2h) for low and high stability samples must be within ± 30.0% of control concentration (T0h).
- Precision: The CV of the stability samples should not exceed 30.0%.
- A minimum of 2/3 of the replicates must have results that can be evaluated.

Assessment at MQC level at room temperature for 2 hours prepared with K_2_ EDTA failed in run 48 due to high %CV and repeated in run 50.

No significant interference was observed in the whole blood. Stability was demonstrated.

#### Test item

The test item is identical with the reference item.

#### Reference item

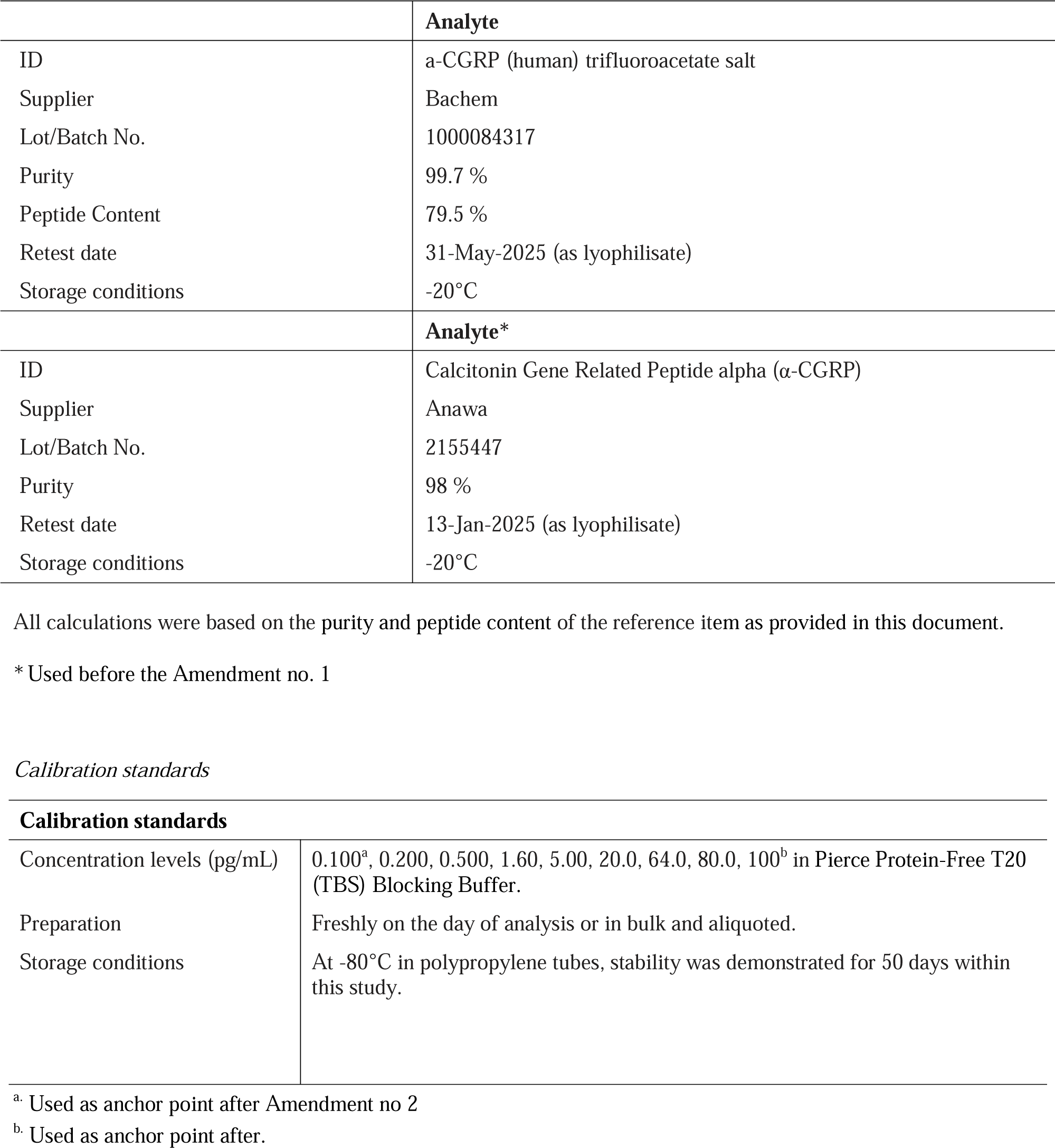

|  | Analyte |
| --- | --- |
| ID | a-CGRP (human) trifluoroacetate salt |
| Supplier | Bachem |
| Lot/Batch No. | 1000084317 |
| Purity | 99.7 % |
| Peptide Content | 79.5 % |
| Retest date | 31-May-2025 (as lyophilisate) |
| Storage conditions | -20°C |
|  | Analyte* |
| ID | Calcitonin Gene Related Peptide alpha (α-CGRP) |
| Supplier | Anawa |
| Lot/Batch No. | 2155447 |
| Purity | 98 % |
| Retest date | 13-Jan-2025 (as lyophilisate) |
| Storage conditions | -20°C |

| Calibration standards |  |
| --- | --- |
| Concentration levels (pg/mL) | 0.100<sup>a</sup>, 0.200, 0.500, 1.60, 5.00, 20.0, 64.0, 80.0, 100<sup>b</sup> in Pierce Protein-Free T20 (TBS) Blocking Buffer. |
| Preparation | Freshly on the day of analysis or in bulk and aliquoted. |
| Storage conditions | At -80°C in polypropylene tubes, stability was demonstrated for 50 days within this study. |

<sup>a</sup>. Used as anchor point after Amendment no 2
<sup>b</sup>. Used as anchor point after.

## RESULTS

### PACAP Bioanalysis - Rodents

To determine utility of the PACAP MSD S-PLEX assay for measurement of circulating PACAP levels in rodents, we collected plasma from vehicle and PACAP38-challenged mice and measured PACAP levels.

Circulating PACAP in vehicle-treated control mice was estimated to be 2,296 fg/ml, whereas in the presence of inhibitor, it was 2,734 fg/ml (**Figure 3**). This suggests that the presence of DPP-IV inhibitor, only nominally limited degradation of endogenous PACAP. Circulating PACAP in PACAP-challenged mice was estimated to be 494,295 fg/ml, whereas in the presence of inhibitor, it was 478,262 fg/ml (**Figure 3**). This suggests that the presence of DPP-IV inhibitor had no marked effect on the degradation of exogenously administered PACAP. These data confirm that our highly sensitive PACAP assay can be used to measure circulating levels of both endogenous and administrated PACAP38 in mouse plasma. In these samples, the vehicle treated mice had lower PACAP levels compared to PACAP38-challengedmice (**Figure 3**). Levels varied between individual mice in both groups and three samples treated with PACAP38 were measured above the upper limit of quantification. Addition of a DPP-IV inhibitor in the tube after collection of plasma did not improve the stability of the PACAP peptide.

**Figure 3.**
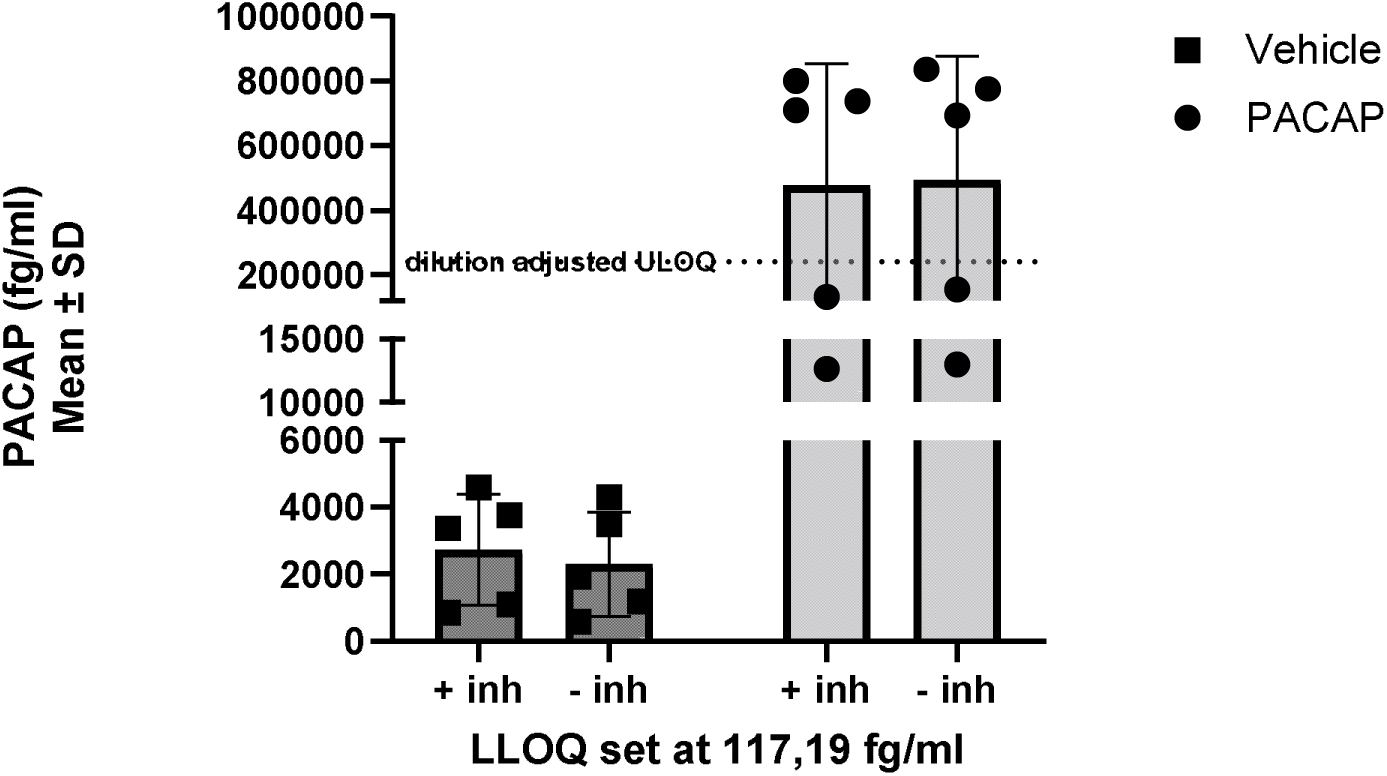
Circulating PACAP level in control and PACAP-challenged C57BL6 mice. Figure shows graph of circulating PACAP level in control and PACAP-challenged C57BL6 mice.No significant differences between the groups were observed. Mean values ± SD are shown (n = 5).

We also collected plasma from control and PACAP38-challenged rats to determine utility of the PACAP MSD S-PLEX assay for measurement of circulating PACAP levels in rat plasma. As before, the plasma samples were collected in the presence or absence of DPP-IV inhibitor, to limit degradation of endogenous PACAP. Circulating PACAP in vehicle-treated control rats was estimated to be 1,425.45 fg/ml, whereas in the presence of inhibitor, it was 1,919 fg/ml (**Figure 4**). This suggests that the presence of DPP-IV inhibitor, only nominally limited degradation of endogenous PACAP, as observed with the mouse samples. Circulating free PACAP in PACAP-challenged rats was estimated to be 24,405.4 fg/ml, whereas in the presence of inhibitor, it was 28,526.8 fg/ml (**Figure 4**). This suggests that the presence of DPP-IV inhibitor had no marked effect on the degradation of exogenously administered PACAP. These data confirm that our highly sensitive PACAP assay can be used to measure circulating levels of endogenous and administrated PACAP38 in rat plasma. In the current experiment, vehicle treated rats had much lower PACAP levels compared to PACAP38-challengedrats, and levels varied between individual rats in both groups. As with the mouse samples, addition of a DPP-IV inhibitor in the tube after collection of plasma did not improve the stability of the PACAP peptide.

**Figure 4.**
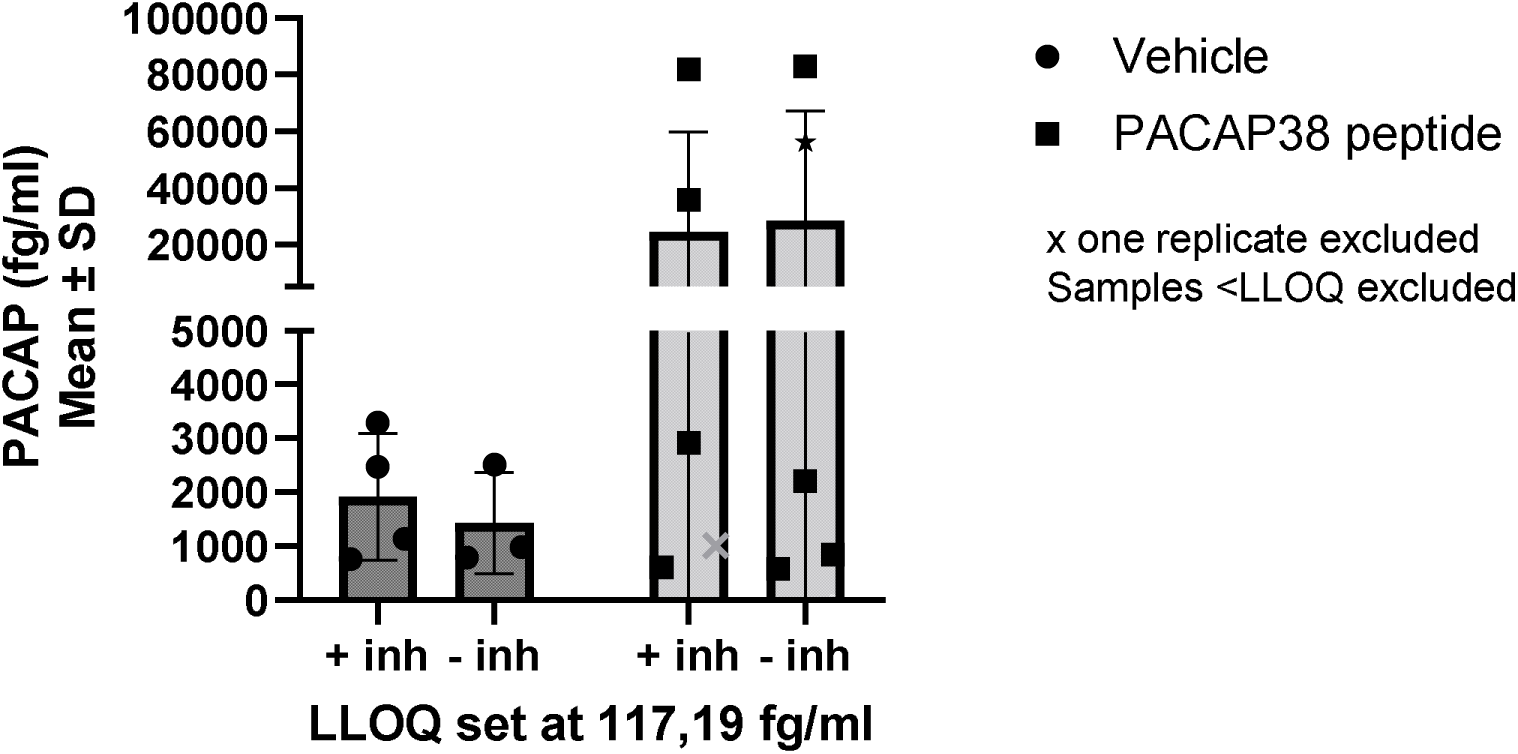
Circulating PACAP level in control and PACAP-challenged SD rats. Figure shows graph of circulating PACAP level in Control and PACAP-challenged SD rats. Nono significant differences between the groups were found. Mean values ± SD are shown (n = 5).

### PACAP Bioanalysis – Human Samples

As a potential migraine biomarker, the stability of PACAP peptide under ex vivo conditions is critical for their clinical applications, therefore we aimed to determine whether there was a more optimal way to stabilize circulating PACAP, and the potential impact of instability in blood on measurements of circulating PACAP levels. To limit degradation of endogenous PACAP, blood from 24 healthy donors was collected directly into P800 or EDTA tubes. After plasma collection, samples were stored at −80°C for 3 months prior to analysis. The circulating plasma PACAP levels in the individual healthy donors were variable (**Figure 5**), and the use of P800 tubes resulted in slightly higher mean concentrations of PACAP compared to EDTA tubes (**Figure 5**). This suggest that clinical samples should optimally be collected in P800 tubes for peptide analysis. The methods do seem to perform more robustly in P800 plasma (F/T stability, assay sensitivity etc) However, the data generated in normal EDTA tubes correlate with the data generated in P800 tubes. It is still unknown what impact the longer-term storage in EDTA tubes over P800 could have on the circulating PACAP levels, as quantified in the PACAP MSD S-PLEX assay. It may be that, if the samples will be analysed within a short time (< 6 months) after sampling the loss of integrity may be limited.

**Figure 5.**
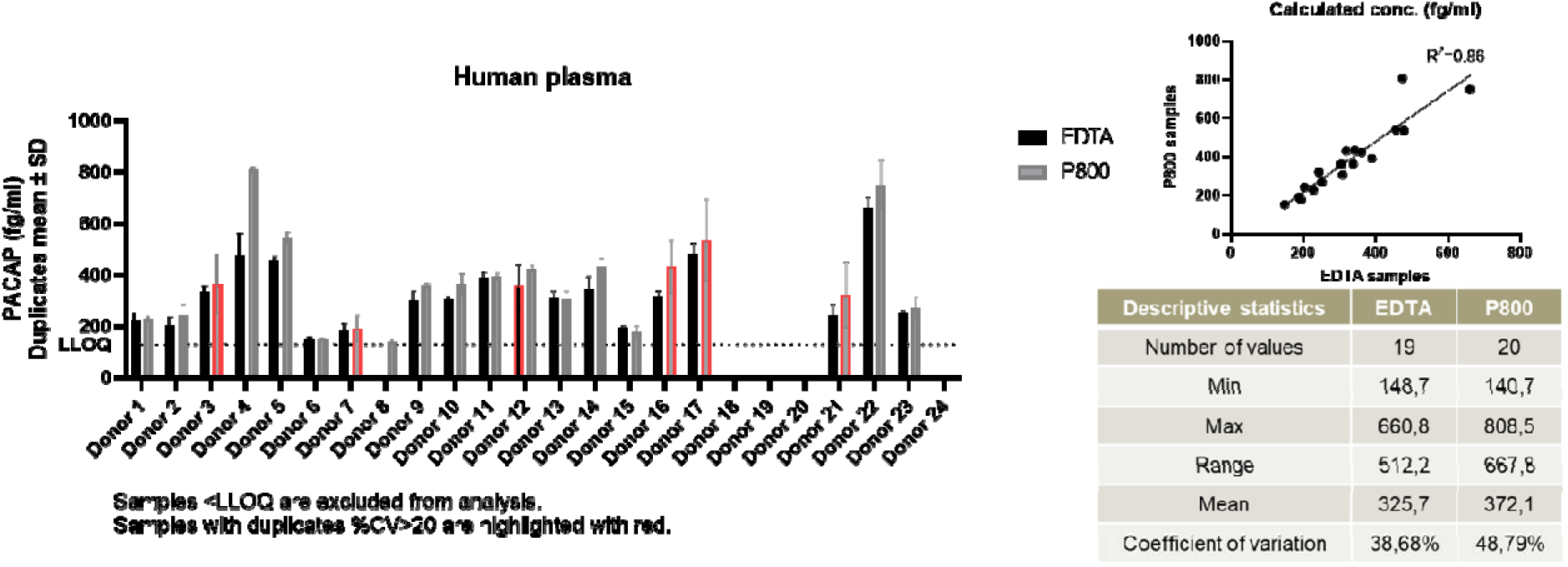
Circulating PACAP level in Healthy donor plasma samples. Figure shows graph of circulating PACAP level in human plasma samples (n = 24). Both the P800 stabilized plasma and EDTA plasma were analyzed in duplicates in the PACAP MSD S-PLEX assay after 1:2 dilution. Mean values ± SD are shown.

Next, we prospectively collected plasma from people with migraine and matched healthy control samples in regular EDTA tubes, to mimic standard practice, and baseline levels for PACAP was measured. The mean total amount of PACAP in healthy control subjects was approximately 190.5 fg/ml (**Figure 6**), while it was 203.7 fg/ml in people with migraine (**Figure 6**). There was no significant difference between PACAP levels in migraine subjects versus levels in healthy controls.

**Figure 6.**
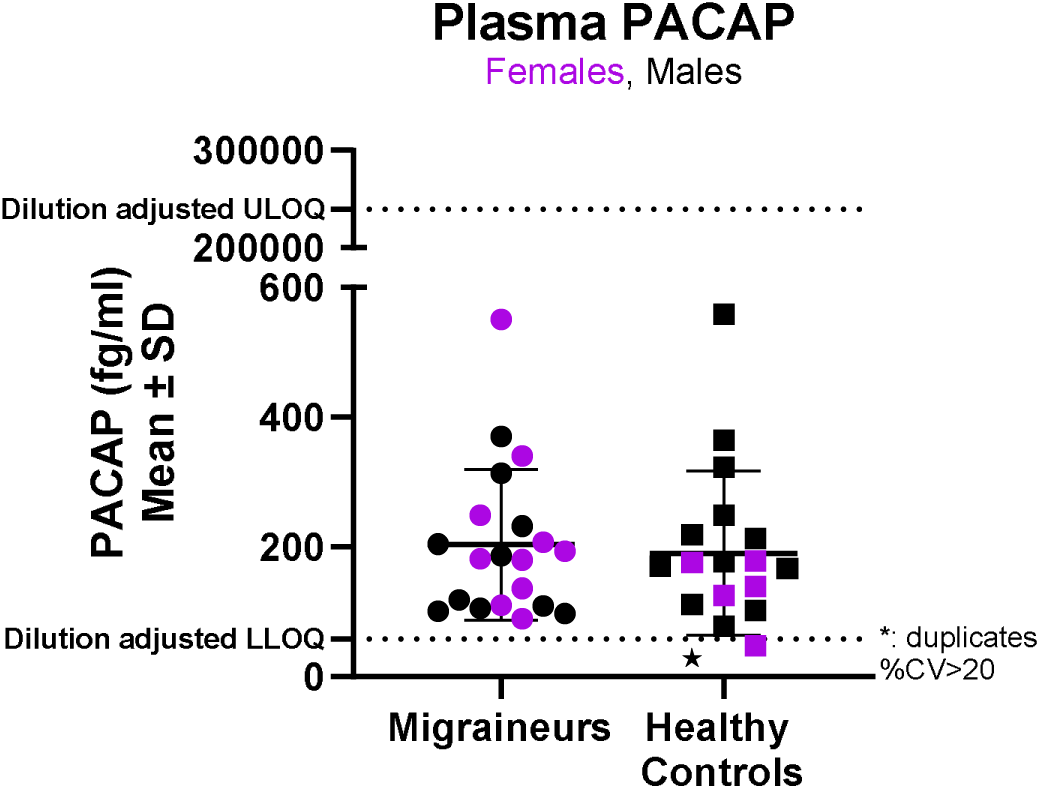
Baseline levels of circulating PACAP level in healthy controls and people with migraine Figure shows graph of circulating plasma pituitary adenylate cyclase-activating polypeptide (PACAP) levels in the migraine and healthy control groups. 40 human plasma samples were analyzed in duplicates in the PACAP MSD S-PLEX assay after 1:2 dilution. Each block represents the mean ± SD values of the results. Plasma levels of these peptides between Migraine (n = 20) and control (n = 20) groups are compared by independent samples t-test or Mann-Whitney U test. Mean values ± SD are shown.

We followed up this analysis by prospectively collecting a new set of samples from people with migraine during and after and attack period, together with a new set of samples from healthy controls. As before, the mean circulating levels of PACAP (226.86 fg/ml PACAP) was variable in the heathy controls (**Figure 7**) but was consistent with previously measured levels (**Figure 6**). The levels of PACAP during a migraine attack period were shown to be higher (495.2 fg/ml PACAP) but markedly reduced outside the attack period (213.7 fg/ml PACAP) (**Figure 7**). However, 6 out of 12 Migraineurs showed increased circulating plasma PACAP levels during their attack period compared to post attack. The average PACAP levels of the post Migraine attack samples were similar to the average level of the Healthy Control subjects (226.86 fg/ml versus 213.7 fg/ml).

**Figure 7.**
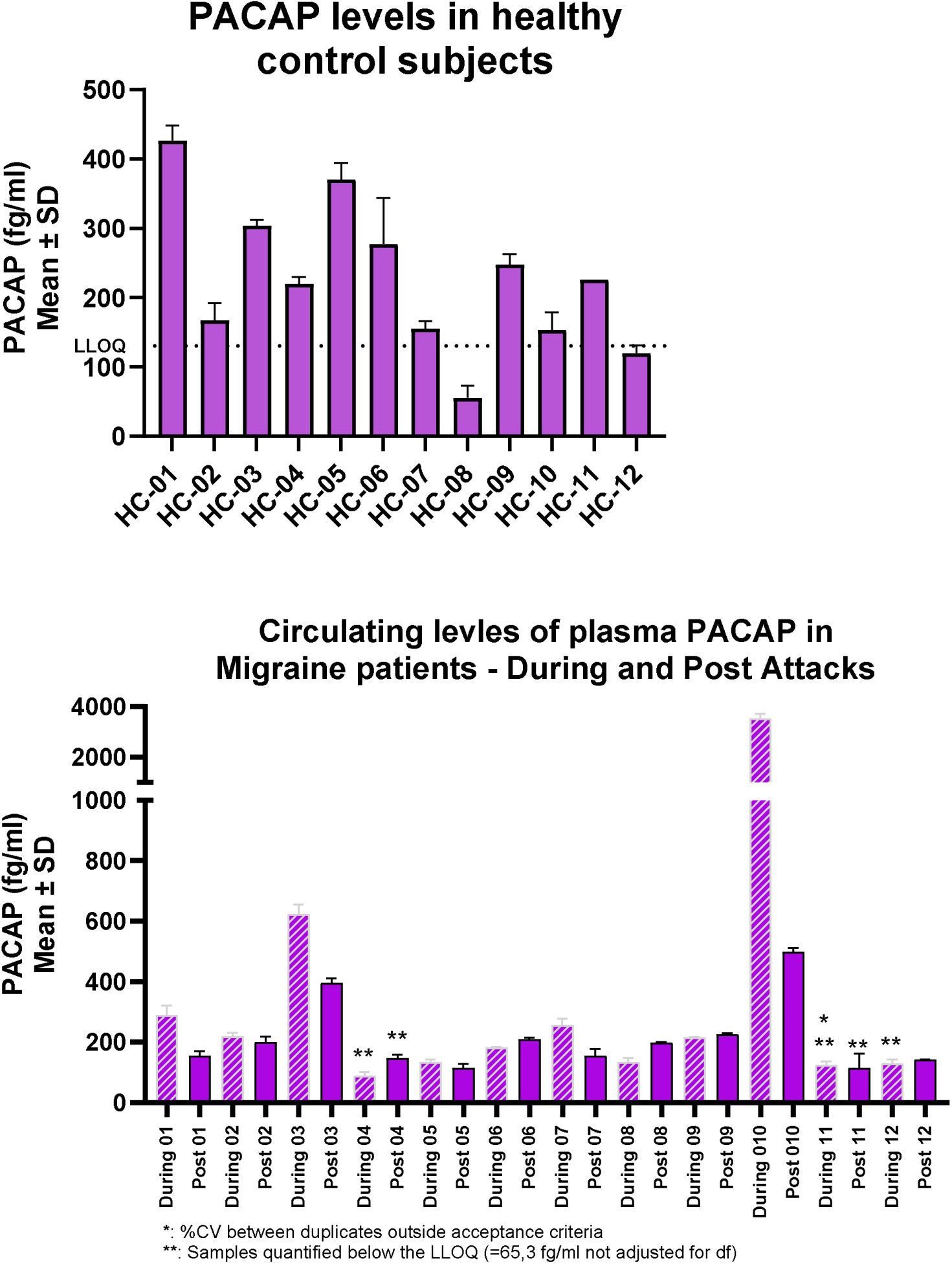
Circulating PACAP levels in healthy controls and people with migraine during and post migraine attacks Figure shows graph of circulating PACAP levels in the healthy control and migraine groups.36 human plasma samples were analyzed in duplicates in the PACAP MSD S-PLEX assay after 1:2 dilution. Plasma levels of these peptides between migraine (ictal and interictal, n = 38) were compared by independent samples t-test or Mann-Whitney U test. n.s.: non-significance (not shown here).

### CGRP Bioanalysis – Human Samples

Baseline levels for CGRP in plasma from people with migraine were determined and compared to values obtained from healthy controls. The mean total amount of CGRP in control subjects was approximately 0.708 pg/ml (**Figure 8**), while it was 0.658 pg/ml in people with migraine (**Figure 8**). There was no significant difference between CGRP levels in migraine subjects versus levels in healthy controls.

**Figure 8.**
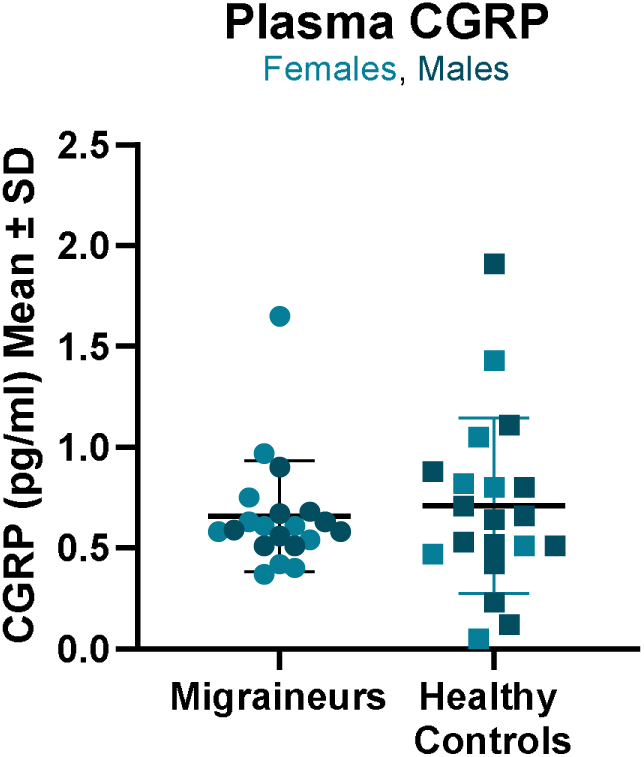
Baseline levels of circulating PACAP level in healthy controls and people with Migraine Figure shows graph of circulating plasma calcitonin gene-related peptide (CGRP) levels in the migraine and healthy control groups. 40 human plasma samples were analyzed in duplicates in the CGRP MSD S-PLEX assay MSD after 1:4 dilution. Each block represents the mean ± SD values of the results. Plasma levels of these peptides between migraine (n = 20) and control (n = 20) groups are compared by independent samples t-test or Mann-Whitney U test. Mean values ± SD are shown.

We followed up this analysis by measuring CGRP levels in the prospective samples where we previously measured PACAP. In those samples, the circulating levels of CGRP (0.87 pg/ml) in the heathy controls was not variable (**Figure 9**) but was consistent with previously measured levels (**Figure 8**). As shown in (**Figure 9**), we observed that like the profile of PACAP, the levels of circulating CGRP during a migraine attack period were variable and shown to be higher (2.06 pg/ml) and reduced outside the attack period (1.04 pg/ml). Circulating plasma CGRP levels in people with migraine were consistent with previously measured levels (**Figure 8**). However, 8 out of 12 people with migraine showed increased circulating plasma CGRP levels during the attack period compared to post attack.

**Figure 9.**
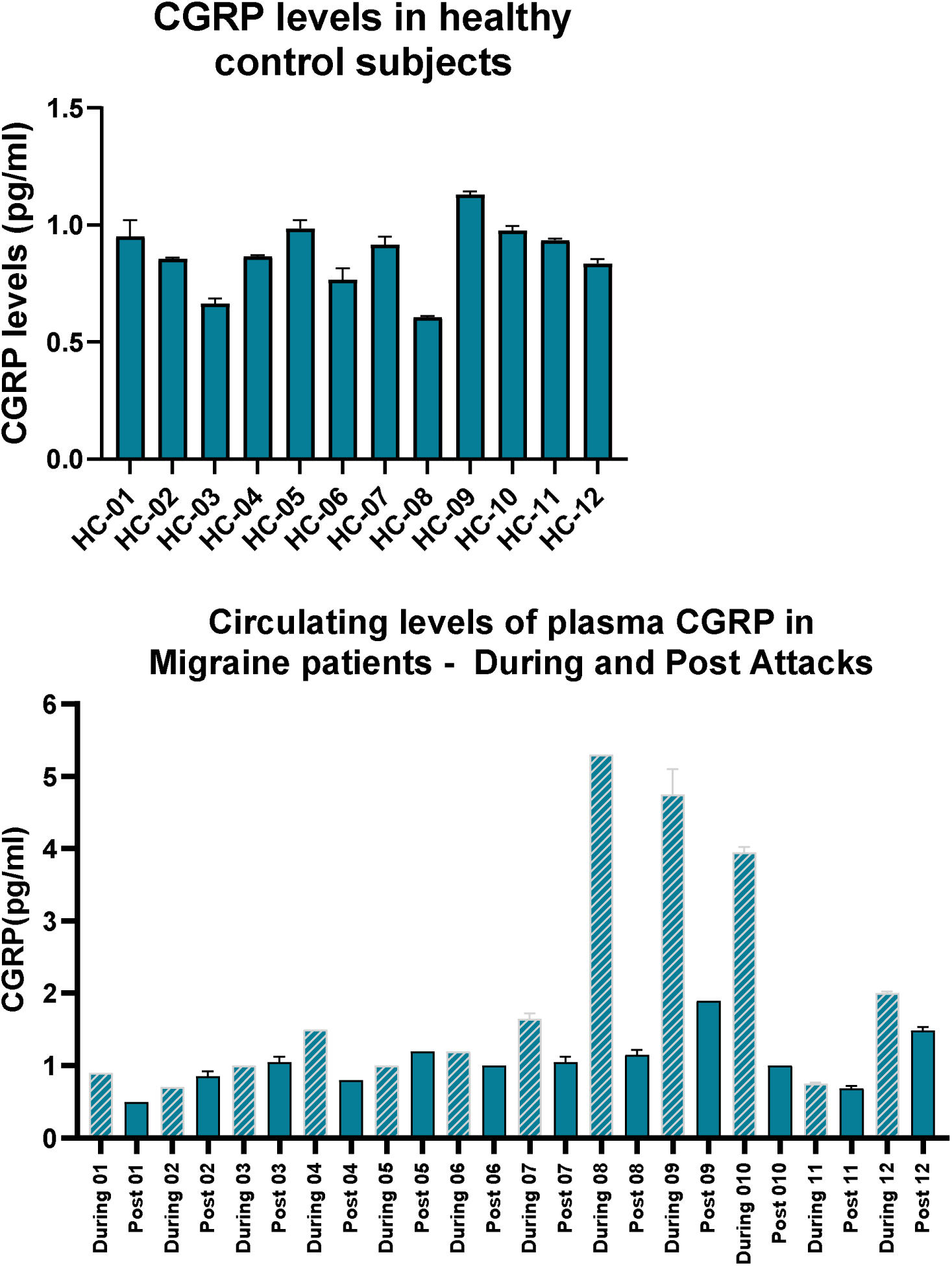
Circulating CGRP levels in healthy controls and people with migraine during and post migraine attacks Figure shows graph of circulating CGRP levels in the migraine and healthy control groups. 36 human plasma samples were analysed in duplicates in the CGRP MSD S-PLEX assay after 1:4 dilution. Plasma levels of these peptides between Migraine (ictal and interictal, n = 36) were compared by independent samples t-test or Mann-Whitney U test. n.s.: non-significance (not shown here).

### CGRP and PACAP assay method validation

The bioanalytical method met the requirements for the tested validation assessments and was fit for purpose for exploring CGRP and PACAP concentrations in human plasma. Sampling in protease-stabilised tubes is recommended. Long-term stability assessments are ongoing in a separate study.

## DISCUSSION

The neuropeptides CGRP and PACAP have been recognized as key players in migraine pathophysiology. This has driven the development of mechanism-based medicines for preventive treatment of migraine. Accurate and precise quantification of the circulating neuropeptides is essential to the continued development of these mechanism-based medicines. In addition, they may be beneficial for patient selection in clinical trials, to demonstrate treatment effects of drugs that specifically target these peptides and for clinical assessment of the utility of these neuropeptides as biomarkers of specific indications (people with Migraine) or patient subgroup of an indication (subgroup of people with Intrusive-Hypervigilant PTSD have elevated PACAP compared to people with PTSD). ^25^. In that regard, we have developed highly sensitive assays for measuring PACAP and CGRP, and we have successfully used the assays to measure CGRP and PACAP in mice, rats, and human samples.

These data demonstrate that baseline levels of CGRP and PACAP between attacks are coordinately elevated in some people with migraine, but the precise temporal profile of such changes likely requires a more thorough longitudinal profiling study. There was no significant difference between PACAP levels in migraine subjects versus levels in healthy controls, and this was in contrast to some prior published observations ^26^, but similar to published data on serum PACAP levels in people with migraine ^27^. There was also no significant difference between CGRP levels in migraine subjects versus levels in healthy controls, and this was similar to some prior published observations ^26^. Our data also suggest that the same peptide may not be elevated at the same time in the same person, which is consistent to the heterogeneity of migraine, and potentially also migraine triggers.

Overall, the measurement of PACAP and CGRP in mouse, rat and human samples supports the utility of the highly sensitive assays we have developed and formed the basis for validating these assays for clinical use. Direct measurement of low abundance target protein presents numerous analytical challenges, in particular, sensitivity, uncertainty about their binding partners, turnover rates and breakdown products^28^. Thus, there remains a need for development of orthogonal assays (LC-MS) that may allow us to circumvent these challenges. While our assays provide sensitive detection, the development of LC-MS assays could offer the added advantage of distinguishing specific peptide lengths, enabling precise characterization of degradation products that could play a role in the pathophysiology of Migraine.

## Data Availability

All data produced in the present study are available upon reasonable request to the authors

## Acknowledgments

Development of the assays was funded by H. Lundbeck A/S (Copenhagen, Denmark)

